# Feasibility of Volumetric Analysis using Bedside Ultra-Low-Field Portable Magnetic Resonance Imaging in Patients receiving Extracorporeal Membrane Oxygenation

**DOI:** 10.64898/2026.04.09.26350481

**Authors:** Melissa D. Stockbridge, Andreia V. Faria, Voss Neal, Isidora Diaz-Carr, Zoe Soulé, Yaman B. Ahmad, Shivalika Khanduja, Glenn Whitman, Argye E. Hillis, Sung-Min Cho

## Abstract

The SAFE MRI ECMO (NCT05469139) study established the safety of ultra-low-field 64mT MRI in patients receiving extracorporeal membrane oxygenation (ECMO) in the setting of intensive care and demonstrated that these images were highly sensitive in detecting acquired brain injuries. This retrospective analysis of prospectively collected observational data sought to expand on these findings in light of the crucial need for neurological monitoring while patients receive ECMO by evaluating the feasibility of volumetric analyses derived from ultra-low-field MR images. T2-weighted scans from thirty patients who received ultra-low-field MRI while undergoing ECMO at Johns Hopkins Hospital were analyzed using a volumetric pipeline to determine whole brain volume and volumes of total grey matter, total white matter, subcortical grey matter, ventricles, left hemisphere, right hemisphere, telencephalon, left and right lateral ventricles, the total intracranial volume, and the cerebellum. Segmented brain volumes in patients undergoing ECMO were comparable to measurements obtained using conventional field and ultra-low-field MRI in the absence of ECMO instrumentation. The subgroup analysis demonstrated subtle volumetric differences between patients supported with venoarterial ECMO and those receiving venovenous ECMO. These data provide the first evidence that ultra-low-field MRI provides volumetric measurements comparable to conventional field-strength MRI, even in the presence of ECMO circuitry, supporting its feasibility for neuroimaging in critically ill patients.

## Introduction

Decreases in regional and whole brain volume are important imaging-based biomarkers of disease, including neurodegenerative diseases [1-6], and psychiatric illnesses [7-11]. Conventional magnetic resonance imaging (MRI) is the gold standard for assessing brain volume, due to improved soft tissue contrast supporting high-accuracy tissue segmentation and better spatial resolution [12-14]. However, MR technology has numerous limitations, both for institutions and individuals. Many institutions cannot afford the substantial upfront costs associated with purchasing and maintaining large, expensive high-field MR scanners, nor the dedicated infrastructure or specialized staffing required to operate them. There is also the dependence on non-renewable helium, which recently became especially problematic following global helium shortages and dramatic price increases [15], with likely future cost fluctuations further limiting sustainability and access. Though fewer limitations exist today than in years past, some individuals with implanted devices still cannot safely receive MRI, and prolonged image acquisition times result in a high risk of poor image quality due to patient movement and non-compliance.

Patients receiving extracorporeal membrane oxygenation (ECMO) represent a unique class of patients for whom MRI can provide great clinical value. There is an extensive literature emphasizing the need for neurological monitoring while patients receive ECMO, as it is associated with an increased risk of acute brain injury [16-19], leading to poorer outcomes [20]. Neuroimaging patients receiving ECMO presents unique challenges. Due to the severity of illness, transporting these patients to radiology suites is often challenging with potential adverse events [21]. Even when transport is feasible, the 1.5–3 Tesla magnetic fields required for conventional clinical MRI raise significant concerns regarding heating, migration, and potential malfunction of ECMO equipment [22].

Recent innovative work examined the feasibility and safety of using ultra-low-field portable MRI (pMRI) to acquire neuroimages in ECMO patients [22, 23]. A multicenter cohort of 64 ECMO patients found bedside pMRI demonstrated high sensitivity for detecting ABI, identifying intracranial hemorrhage, cerebral edema, and ischemia with diagnostic accuracy comparable to, and in some cases exceeding, head CT. In addition to producing comparable images to 1.5 or 3T T1- and T2-weighed scans, these scanners are less expensive to purchase and run, do not require helium or advance infrastructure, and generate only a 64 mT field, 160 times weaker than that of a conventional scanner. However, the full breadth of how 64 mT images may be clinically informative has yet to be fully characterized, and reduced image resolution and artifacts in pMRI have been problematic in certain applications of conventional neuroimage processing pipelines [24].

Beyond the detection of acute brain injury, 64 mT acquisitions may also enable quantitative measurement of brain volume in adult ECMO patients. Brain volume decreases over time are gradual in the typical aging population. Beginning around age 40, normal adults begin losing brain volume and the rate slowly accelerates year over year [25, 26]. Multiple neurodegenerative diseases are associated with faster rates of volume loss [27]. *Accelerated* brain atrophy and white matter lesions have been observed in association with intensive care unit (ICU) hospitalizations [28-30]. A large, retrospective cohort study found that ICU admission was associated with significant accelerated temporal, frontal, and parietal cortical thinning and declines in volume in the hippocampus and corpus callosum [31]. ICU admission was not associated with increased cortical amyloid or thinning in the entorhinal cortex, inferior temporal cortex, middle temporal cortex, and fusiform, as is commonly associated with Alzheimer’s disease pathology [32]. Rapidly progressing brain atrophy also was observed in a sample of 84 adult patients who received mechanical ventilation for a median of 30 days [33]. Among those for whom atrophy was observed, an average pre-post ventilation brain volume decrease of 3.3 ± 2.4% was observed (a significant change).

Despite the potential value of volumetric assessments in this context, a key limitation previously identified in the literature has been the difficulty in segmentation for volumetric analysis when done directly from low-field MRI images [24]. The aim of this work was to determine the feasibility of volumetric analyses using 64 mT MRI in patients receiving ECMO. Structural images were processed with an optimized pipeline for low-field acquisitions to determine if additional sources of artifact preclude this application.

## Materials and Methods

### Participants

In this retrospective analysis of prospectively collected data from 30 patients who received pMRI from August 2022 to November 2023 while undergoing ECMO at Johns Hopkins Hospital as part of the SAFE MRI ECMO study [NCT05469139; 22]. Included patients were adults undergoing either venoarterial (VA ECMO) or venovenous ECMO (VV ECMO). Exclusion criteria followed the SAFE MRI ECMO protocol, which used a modified version of standard MRI contraindications. Namely, Swan-Ganz catheters [34] and intra-aortic ballon pumps [35], devices traditionally considered unsafe in 3T scanners, were permitted as their safety in 64 mT pMRI had been previously established.

### Image acquisition

A comprehensive description of the methods employed is present in the definitive record results cited above. In summary, 64 mT MRIs were performed in the patient’s own bed using a Swoop MRI system (Hyperfine, Inc, Guilford, CT; hardware versions 1.6 and 1.8 with software versions 8.6.1 and 8.7), and vital signs were monitored continuously throughout the scans. T1-weighted, T2-weighted, fluid-attenuated inversion recovery (FLAIR), and diffusion-weighted imaging (DWI) with automatically calculated apparent diffusion coefficient (ADC) map sequences were acquired.

### Image processing

All processing was completed in FreeSurfer 8 [36, 37]. Freesurfer morphometric procedures previously have been demonstrated to show good test-retest reliability across scanner manufacturers and across field strengths [38, 39]. Segmentation of the T2-weighted images was completed using the SynthSeg function [40, 41] in all samples, for consistency.

In anticipation of the present research, test-retest reproducibility of this volumetric pipeline was affirmed in 10 neurologically typical, English-speaking adult volunteers as a contemporaneous methodological check using Swoop MRI system software version 8.8.1 [42]. Eighteen individuals from the existing NITRC Multi-Modal MRI Reproducibility Resource who underwent repeated 3T scans were utilized as a comparator [43]. Excellent test-retest stability [44] was observed in interclass correlation coefficients between repeated volume measurements of total intracerebral volume, total grey matter, subcortical gray matter, and white matter, left and right hemispheres separately, total ventricles, left and right lateral ventricles separately and telencephalon derived from T2-weighted scans acquired on 64mT and 3T scanners (ICC >= 0.9).

Greater variability was seen in cerebellar volumes measured from 64 mT (ICC = 0.73) compared with 3 T scanners (ICC = 0.99). The brain volumes were consistent with known normative growth curves for whole brain volumes of large populations (nearly 100,000 individuals) in BrainChart.io [45]. All work was conducted following ethical approval by the Johns Hopkins University School of Medicine Institutional Review Board: Federal Wide Assurance # FWA00005752-JHUSOM; IRB00216321 “Retrospective Analysis of Outcomes of Patients on Extracorporeal Membrane Oxygenation,” IRB-6 initially approved 10/22/2019; NA_00042097 “Neural Basis of Language and Cognitive Deficits in Acute Stroke and Recovery,” IRB-2 initially approved 10/21/2010 (neurologically normal controls); IRB00501130 “Advanced Imaging of Primary Progressive Aphasia,” IRB-2 initially approved 6/26/2025 (neurologically normal controls). Patients or their legally authorized representatives provided informed consent. Data from IRB00216321 were de-identified, while data from NA_00042097 and IRB00501130, which are ongoing observational investigations, were available to co-investigators.

### Analysis

Data were accessed November 1, 2025. The measurements derived from T2-weighted scans acquired during ECMO were calculated for the volume of total grey matter, total white matter, subcortical grey matter, ventricles, left hemisphere, right hemisphere, telencephalon, left and right lateral ventricles, the total intracranial volume, and the cerebellum using FreeSurfer. Patients receiving VA and VV ECMO were considered separately for exploratory comparison.

The brain volumes from ECMO were compared to those from healthy volunteers on pMRI and 3T test-retest sets and plotted on known normative growth curves for whole brain volume of large populations (nearly 100,000 individuals) in BrainChart.io [45] to check whether these volumes fall within the normal population range for the same age and sex. Groups were not statistically compared, as the available samples were not age-paired groups.

## Results

### ECMO cohort characteristics

Of the 34 patients receiving ECMO included in the study, 16 were male and 18 were female, as summarized in Table 1. Median age was similar between sexes, with males aged 56.5 years and females aged 58 years. Median body mass index was 28.7 kg/m^2^ in males and 30.7 kg/m^2^ in females.

**Table 1.** Participant characteristics.

|  | Male (n=16) | Female (n=18) |
| --- | --- | --- |
| **Demographics** |  |  |
| Age (years) | 57 (42–68) | 58 (46–70) |
| Body mass index (kg/m<sup>2</sup>) | 28.7 (26.9–34.3) | 30.7 (28.2–35.3) |
| Race and Ethnicity, n (%) |  |  |
| Asian | 10 (62.5%) | 5 (27.8%) |
| Black | 2 (12.5%) | 10 (55.6%) |
| Hispanic | 2 (12.5%) | 1 (5.6%) |
| Other | 2 (12.5%) | 1 (5.6%) |
| White | 0 (0%) | 1 (5.6%) |
| **Medical History, n (%)** |  |  |
| Congestive heart failure | 6 (37.5%) | 5 (27.8%) |
| Chronic kidney disease | 4 (25.0%) | 4 (22.2%) |
| Atrial fibrillation | 2 (12.5%) | 5 (27.8%) |
| **Pre-ECMO variables** |  |  |
| Glasgow Coma Scale score | 3 (3–10) | 5 (3–6) |
| SOFA score (day 1) | 13 (13–16) | 12 (11–15) |
| Inotrope or vasopressor support, n (%) | 12 (75.0%) | 15 (83.3%) |
| Cardiac arrest, n (%) | 3 (18.8%) | 1 (5.6%) |
| **Pre-MRI Blood Gas\*** |  |  |
| pH | 7.4 (7.4–7.5) | 7.4 (7.4–7.5) |
| PaCO<sub>2</sub>, mmHg | 37 (34–46) | 37 (33–41) |
| PaO<sub>2</sub>, mmHg | 96 (77–164) | 126 (82–161) |
| HCO<sub>3</sub><sup>-</sup>, mEq/L | 25 (24–26) | 24 (22–25) |
| SaO<sub>2</sub>, % | 97 (95–99) | 99 (97–99) |
| FiO<sub>2</sub>, % | 65 (48–100) | 60 (50–95) |
| **ECMO Cannulation Strategy, n (%)** |  |  |
| **VA ECMO** |  |  |
| Central cannulation | 7 (43.8%) | 4 (22.2%) |
| Peripheral cannulation | 5 (31.2%) | 9 (50.0%) |
| **VV ECMO** |  |  |
| Double-lumen cannulation | 4 (25.0%) | 5 (27.8%) |
| **ECMO Indications, n (%)** |  |  |
| **VA ECMO** |  |  |
| Cardiogenic shock | 9 (56.2%) | 9 (50.0%) |
| Post-cardiotomy shock | 3 (18.8%) | 4 (22.2%) |
| **VV ECMO** |  |  |
| Acute respiratory distress syndrome | 2 (12.5%) | 2 (11.1%) |
| COVID-19 | 2 (12.5%) | 2 (11.1%) |
| **ECMO management, n (%)** |  |  |
| Anticoagulation during ECMO | 13 (81.2%) | 17 (94.4%) |
| **ECMO outcomes, n (%)** |  |  |
| Mortality | 10 (62.5%) | 13 (72.2%) |
| Acute brain injury (ABI) | 4 (25.0%) | 11 (61.1%) |
| Ischemic stroke | 3 (18.8%) | 10 (55.6%) |
| Intracerebral hemorrhage | 0 (0%) | 1 (5.6%) |
| Subarachnoid hemorrhage | 0 (0%) | 1 (5.6%) |
| Subdural hematoma | 1 (6.2%) | 0 (0%) |
| **ECMO duration, days** |  |  |
| **VA ECMO** | 10 (5–17) | 9 (8–13) |
| **VV ECMO** | 48 (43–53) | 11 (7–28) |

**Abbreviations:** ABI, acute brain injury; ARDS, acute respiratory distress syndrome; BMI, body mass index; COVID-19, coronavirus disease 2019; ECMO, extracorporeal membrane oxygenation; FiO<sub>2</sub>, fraction of inspired oxygen; GCS, Glasgow Coma Scale; HCO<sub>3</sub><sup>-</sup>, bicarbonate; PaCO<sub>2</sub>, partial pressure of arterial carbon dioxide; PaO<sub>2</sub>, partial pressure of arterial oxygen; SaO<sub>2</sub>, arterial oxygen saturation; SOFA, Sequential Organ Failure Assessment; VA, venoarterial; VV, venovenous.
Continuous variables are reported as median (interquartile range), and categorical variables are reported as n (%) using sex-specific denominators.
\*Pre-MRI blood gas variables were collected within 48 hours of ECMO cannulation; the value closest to the MRI scan was recorded.

Baseline characteristics differed by sex. Males were more frequently Asian, whereas females were more frequently Black. Congestive heart failure and chronic kidney disease were common in both groups, while atrial fibrillation was more prevalent among females. Illness severity was high in both sexes, with low Glasgow Coma Scale scores and elevated SOFA scores. Inotropic or vasopressor support was required in most patients, and cardiac arrest prior to cannulation occurred more frequently among males.

Pre-MRI arterial blood gas values obtained within 48 hours of ECMO cannulation were similar between sexes, including pH, PaCO_2_, PaO_2_, bicarbonate, oxygen saturation, and fraction of inspired oxygen. Both VA and VV ECMO configurations were used. Among patients receiving VA ECMO, males more commonly underwent central cannulation, whereas females more frequently underwent peripheral cannulation. The primary indication for VA ECMO was cardiogenic shock, while VV ECMO was used for acute respiratory distress syndrome and COVID-19.

Anticoagulation during ECMO was administered in most patients. In-hospital mortality occurred in 62.5% of males and 72.2% of females. Acute brain injury was observed more frequently among females than males, driven predominantly by ischemic stroke. Hemorrhagic brain injury was uncommon. ECMO duration was similar between sexes for VA ECMO, while longer durations were observed among males receiving VV ECMO.

The analyzed patients receiving ECMO are described in Table 1. Structural volumes are presented in Table 2. The relationship between established growth curves and measurements taken from ECMO patients and healthy controls scanned with ultra-low field MRI, as well as comparable individuals who received traditional 3T MRI are illustrated in Fig. 1. The dispersion in values between the groups was comparable overall (Fig. 2). Bland-Altman plots by region of interest are provided as Supplemental Material.

**Table 2:** Estimated brain volume by structure.

|  | VV ECMO | VA ECMO | Mean Diff |
| --- | --- | --- | --- |
| Grey matter volume | 527.1 ± 59.9 | 511.1 ± 56.5 | -16 |
| White matter volume | 29.7 ± 16.3 | 33.1 ± 19.5 | 3.4 |
| Subcortical grey matter | 1.8 ± 0.6 | 1.5 ± 0.5 | -0.3 |
| Ventricles | 28.2 ± 6.2 | 21.9 ± 10.2 | -6.3 |
| Left hemisphere | 528.2 ± 64.9 | 508.9 ± 56.4 | -19.3 |
| Right hemisphere | 513.7 ± 60.2 | 488.8 ± 55.1 | -24.9 |
| Telencephalon | 1041.9 ± 124.8 | 997.7 ± 110.5 | -44.1 |
| Left Lateral Ventricle | 15.9 ± 8.3 | 18.1 ± 11.2 | 2.1 |
| Right Lateral Ventricle | 15.5 ± 8.4 | 16.5 ± 8.8 | 1.0 |
| Intracranial volume | 1477.8 ± 169.9 | 1413.2 ± 162.1 | -64.5 |
| Cerebellum | 148.1 ± 24.3 | 120.9 ± 42.9 | -27.2 |

Volumes reported in cc. Mean diff: mean difference. df: degrees of freedom. Statistics reported for exploratory understanding only and are not adjusted for multiple comparisons.

**Fig. 1:**
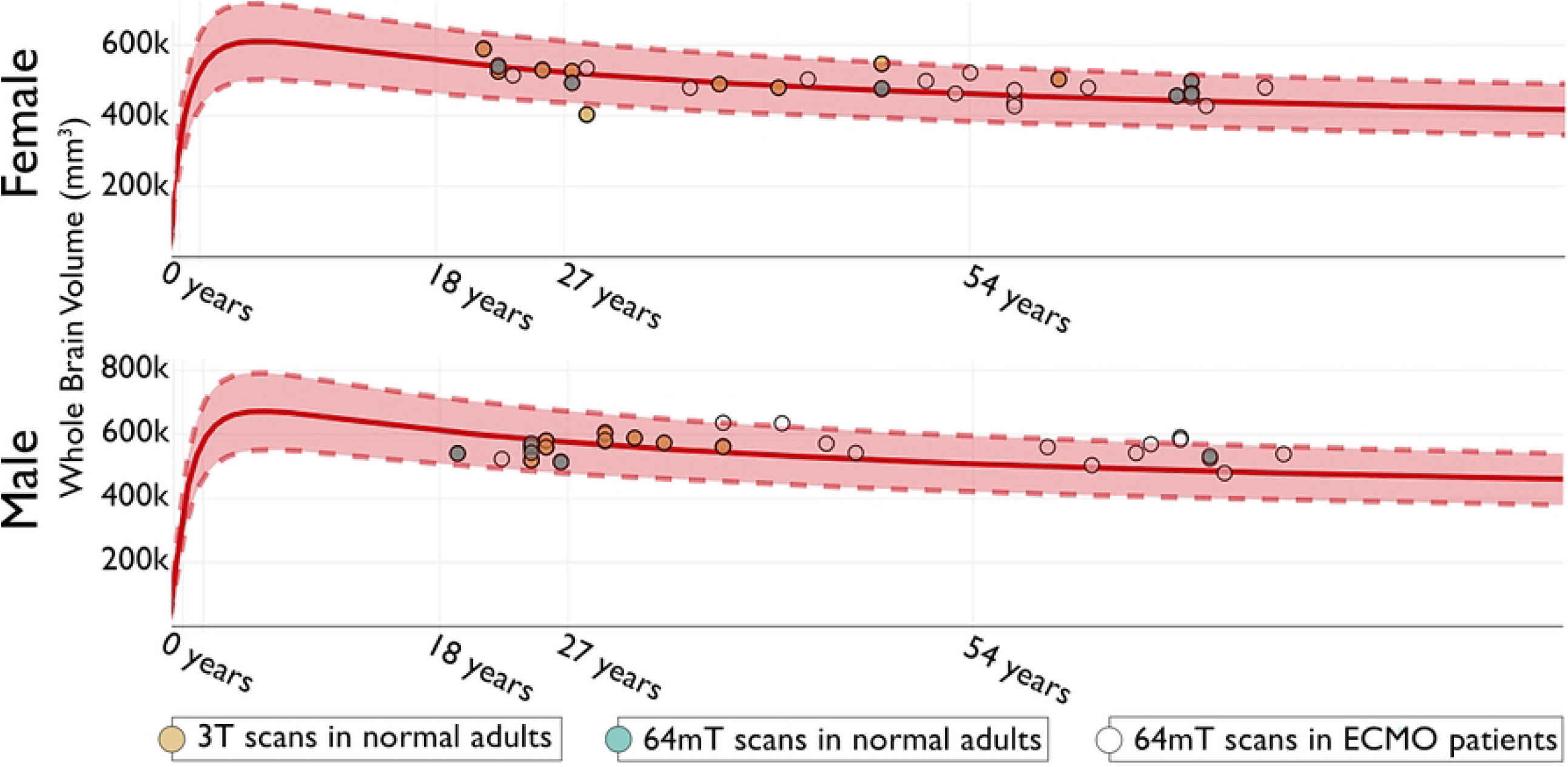
Plot of achieved whole brain volumes among ECMO patients relative to established growth curves. Brain volumes fell within the normal range for individuals of that age and sex. Plots made using BrainChart https://brainchart.shinyapps.io/brainchart/.

**Fig. 2:**
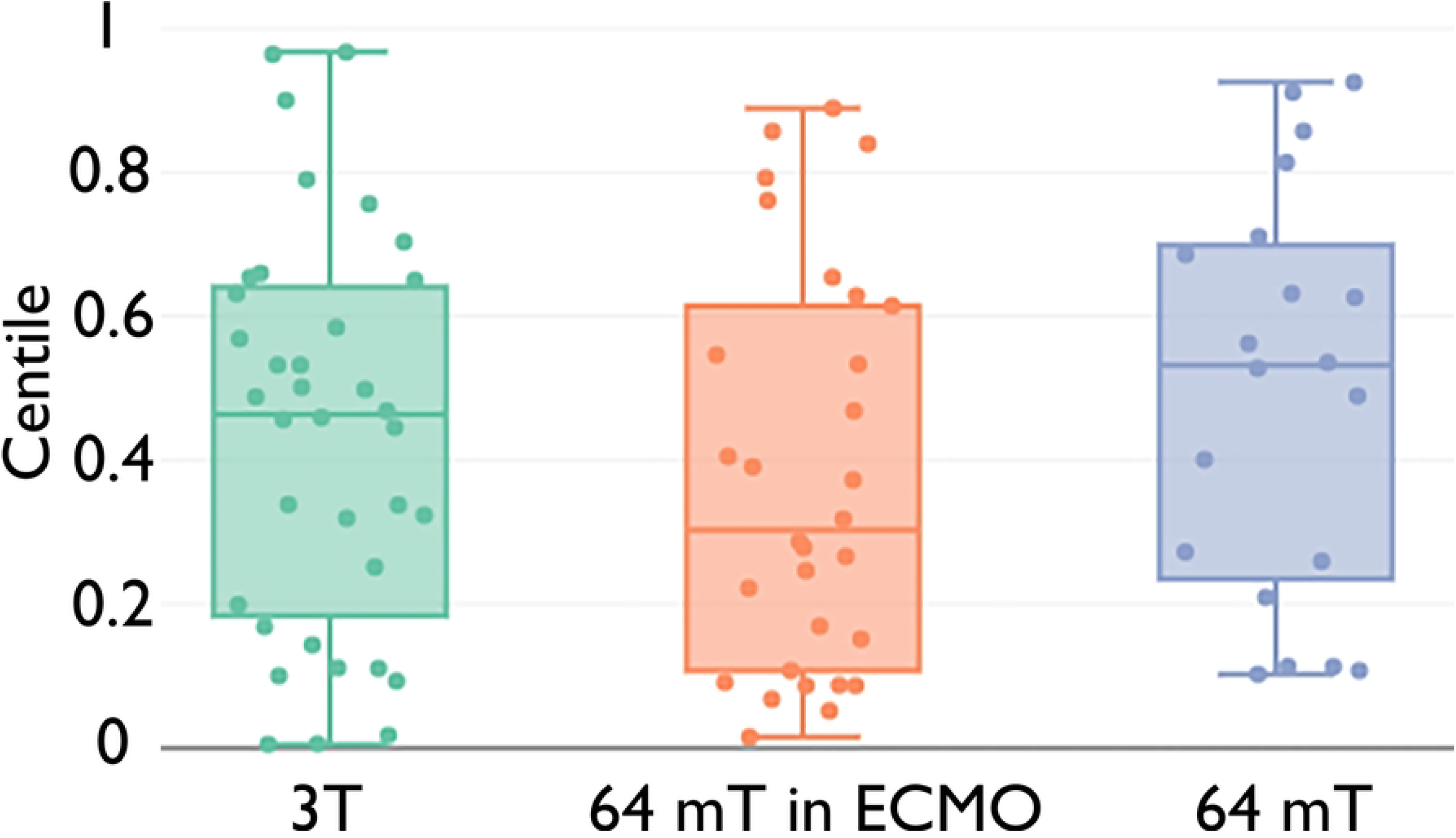
Box plot of whole brain volumes comparing scanner conditions. Patients undergoing ECMO demonstrated slightly lower median volume than neurologically typical patients scanned using 3T or 64mT scanners. Plots made using BrainChart https://brainchart.shinyapps.io/brainchart/.

## Discussion

The aim of the present investigation was to examine the potential clinical utility of low-field pMRI in identifying atypical brain volume loss among patients undergoing ECMO, a potential signal of poorer outcomes [31]. Prior work had identified a limitations of pMRI imaging for volumetric analysis at an earlier phase of technological development [24], which prompted the need to revisit the question in light of recent advances in both hardware and software. Central to the novel reappraisal of volumetric analysis feasibility in pMRI was the introduction of the SynthSeg pipeline [46, 47] and the optimization of that pipeline for use in low-field MRI scans [48].

The results when calculating brain volume in patients undergoing ECMO were comparable to those achieved with conventional MRI and low-field pMRI in the absence of ECMO instrumentation, providing a compelling foundation of evidence that the instruments are not resulting in prohibitive (or even appreciable) signal interference after processing.

Although pursued as a preliminary exploration, subtle differences also appeared to distinguish those who were receiving VA ECMO from those receiving VV ECMO. VV ECMO was associated with larger ventricle and cerebellar volume than VA ECMO. Patients requiring cardiopulmonary support may experience prolonged ventilation, deep sedation, systemic inflammation, and extended critical illness, which may place them at heightened risk for accelerated atrophy. In critically ill populations, delirium has been associated with accelerated brain atrophy over time [31]. In addition, subclinical ischemia, microemboli, impaired autoregulation, and hypoperfusion are well documented in both cardiac surgery and ECMO populations, and postoperative delirium is common, occurring in 26–52% of cardiac surgery patients and strongly associated with long-term cognitive decline [49]. Together, these overlapping factors suggest that neurocognitive vulnerability in cardiopulmonary support populations may reflect not only discrete acute events but also more diffuse, progressive cerebral volume loss. A recent case series supports this concern, documenting the presence of diffuse brain atrophy in neonatal patients receiving ECMO for more than 20 days [50].

Selective cerebellar and brainstem volume loss may reflect posterior circulation vulnerability to altered hemodynamics (low pulsatility [pulse pressure] and more venerability of posterior cerebral autoregulation), particularly loss of pulsatility and non-physiological flow patterns associated with circulatory support. These regions depend on tightly regulated microvascular perfusion and appear less resilient to sustained pulsatility disruption than anterior cortical territories, which is unique in VA ECMO vs. VV ECMO.

## Conclusions

These preliminary results invite future investigation into the utility of concurrent evaluations of whole brain volume change during ECMO. An important area of future investigation will be to determine how more granular structural measurements of volume perform, such as cortical thinning parcellated by lobe as described in Sprung et al. [31], almost certainly requiring a larger number of patients. Prior studies also have focused on the measurement of contralesional hemispheric volume changes in patients with acquired brain injuries to examine these changes over time, and portable technologies present a unique use case for better understanding of the relationship between focal injury and brain volume loss.

In the interval since the completion of the SAFE MRI ECMO study, software and hardware updates have been introduced to improve the quality of acquisitions prior to processing. Example scans utilizing these innovations (Fig. 3) suggest that limitations regarding deep structures and the cerebellum may be lessened in future investigations relying on emerging technologies. Taken together with the present investigation, the findings buoy support for these and other clinical investigations in neurocritical care patients and others whose needs are incompatible with traditional MRI.

**Fig. 3.**
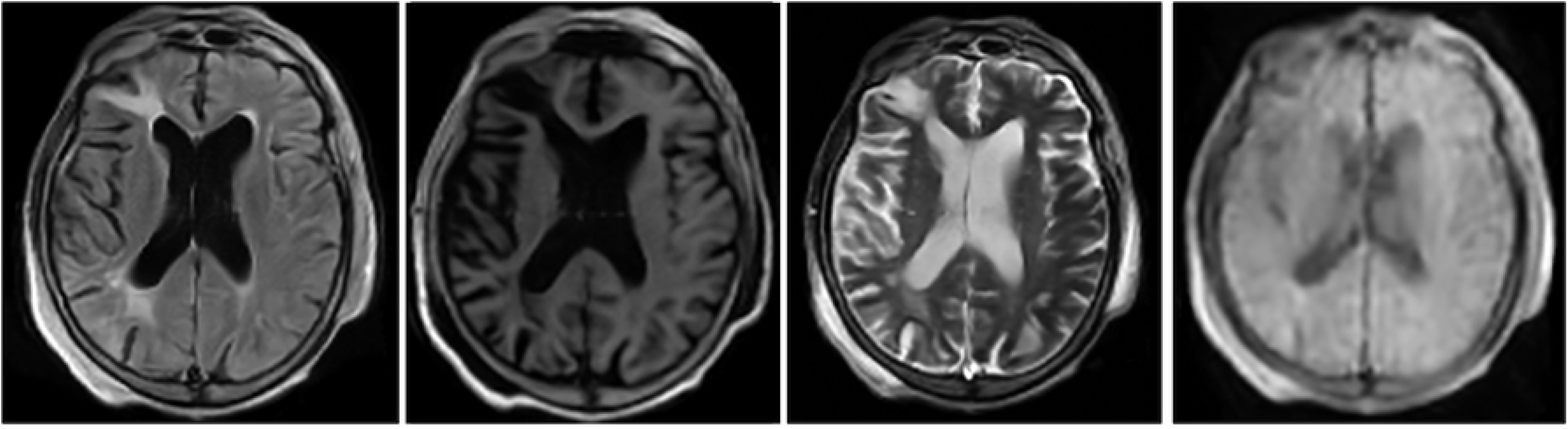
Representative example of portable MRI image quality following software and hardware updates: FLAIR, T1, T2, and diffusion-weighted. Axial brain images acquired using the updated low-field portable MRI platform demonstrate improved signal uniformity and anatomical delineation compared with earlier iterations. Notably, visualization of deeper brain structures and posterior fossa anatomy appears with clarity, suggesting that prior limitations affecting the cerebellum and deep gray matter may be partially mitigated with ongoing technical refinements. These advances support the feasibility of future neuroimaging studies in neurocritical care populations for whom conventional MRI is impractical or contraindicated.

## Data Availability

Data from participants in the original SAFE ECMO study, here used retrospectively, were not subject to a consent form that included the public banking of deidentified data. These data were available from the original study team upon reasonable request, which was granted in the context of IRB00216321 (this analysis). Data from normal controls are available within the NIH-compliant Vivli banked dataset https://doi.org/10.25934/PR00012002.

## Acknowledgments

We gratefully acknowledge the contributions of our participants and their families to the pursuit of clinical research.

